# COVID-19 Prevalence and Trends Among Pregnant and Postpartum Individuals in Maine by Rurality and Pregnancy Conditions

**DOI:** 10.1101/2023.04.21.23288878

**Authors:** Charlie O. Grantham, Christina M. Ackerman-Banks, Heather S. Lipkind, Kristin Palmsten, Katherine A. Ahrens

## Abstract

**Objective:** To estimate COVID-19 diagnosis prevalence and trends among pregnant and postpartum individuals in Maine by rurality and common pregnancy conditions.

**Methods:** We used the Maine Health Data Organization’s All Payer Claims Data to identify deliveries during 2020-2021. We identified COVID-19 during pregnancy (Apr 2020 to Dec 2021 deliveries) and during the first 6 months postpartum (Apr 2020 to Jun 2021 deliveries) using the ICD-10 diagnosis code U071 on medical claims. We used Joinpoint regression software to model trends. We stratified the analysis by rurality of residence (based on ZIP code) and by common pregnancy conditions: gestational diabetes (GDM), hypertensive disorders of pregnancy (HDP), and prenatal depression.

**Results:** We included 13,457 deliveries in our pregnancy and 9,143 deliveries in our postpartum analysis. COVID-19 diagnosis prevalence among pregnant individuals increased from 0.5% in Apr 2020 to 10.5% in Dec 2021 (Oct 2020 was the start of slope [0.43 per month], p<.01). COVID-19 diagnosis prevalence postpartum increased from 0.9% in Apr 2020 to 3.2% in June 2021 deliveries (slope=0.12 per month, p<.01). Trends in prevalence of COVID-19 diagnosis among pregnant individuals living in urban areas were distinct from those living in rural areas (p=.02), with a steeper slope during the first months of the pandemic in urban areas, followed by a later increase among rural residents. Trends among postpartum individuals living in urban areas were distinct from those living in rural areas (p=.03), with a steeper slope for rural residents over the course of the pandemic. Trends in persons with prenatal depression showed a steeper increase in COVID-19 diagnosis prevalence in pregnancy after Dec 2020 (p<.01) and postpartum overall (p<.01) compared to those without prenatal depression. Individuals without GDM and individuals without HDP had steeper increases in COVID-19 diagnosis prevalence in postpartum compared to those without GDM (p<.01) and those without HDP (p=.03).

**Conclusion:** COVID-19 diagnosis among pregnant and postpartum individuals in Maine showed distinct patterns by rurality of residence and select pregnancy conditions. This information can be used for assessing the impact of the COVID-19 pandemic on maternal and infant health.

## INTRODUCTION

SARS-CoV-2 (COVID-19) is a highly transmissible respiratory virus that has resulted in over one million deaths in the U.S. and has been associated with increased risks for adverse perinatal outcomes. Individuals with common pregnancy conditions such as gestational diabetes mellitus (GDM), hypertensive disorders of pregnancy (HPD), and prenatal depression may be at a higher risk of COVID-19 infection and adverse sequelae, though little is known about this topic.^1^ Further, there is a lack of information regarding COVID-19 infection postpartum, even though this time period represents a high-risk time for maternal morbidity and mortality,^2^ and peak infant caregiving.

Pregnant and postpartum individuals living in rural areas may be at greater risk of COVID-19 infection than those living in urban areas.^3^ This could be due lower COVID-19 vaccination rates in some rural areas and lower use of community mitigation strategies, such as policies on masking, closures, and physical distancing.

Therefore, the aim of our study was to estimate the prevalence and trends in COVID-19 diagnosis in persons during pregnancy and in the first 6 months’ postpartum by rurality of residence and common pregnancy conditions. We used data from Maine, the second most rural state in the U.S., where more than 60% of residents live in rural areas.^4^ Compared to the U.S. overall, Maine experienced lower incidence of COVID-19 and had higher vaccination rates for the study period we examined.^5^

## METHODS

We scanned the medical claims of individuals with deliveries (livebirths or stillbirths) identified in the Maine Health Data Organization’s All Payer Claims Data during 2020-2021. We defined COVID-19 diagnosis during pregnancy and during the first 6 months’ postpartum, separately, using the ICD-CM-10 diagnosis code U071 recorded (in any position) on inpatient, outpatient, and professional medical claims. We chose April 2020 as the starting delivery month because it was the first full month after the public health emergency was declared in Maine. We chose June 2021 to define the end of the postpartum cohort because December 2021, which was the last month in our dataset (and beginning of the Omicron wave), marked 6 months’ postpartum. For the postpartum cohort analysis, we restricted this study population to individuals who had continuous health insurance coverage for 6 months’ postpartum. We used Joinpoint regression software to model trends, which allows for several linear trends to be detected and connected together at “joinpoints.” We stratified the analysis by urban and rural residence (based on ZIP code) and by common pregnancy conditions (GDM, HDP, and prenatal depression);^6^ we used tests of coincidence to assess whether trends between groups were identical or not. For delivery months with zero COVID-19 diagnoses found, we imputed case value of 0.1 so the Joinpoint regression model would run.^7^

## RESULTS

### COVID-19 during pregnancy

The prevalence of COVID-19 during pregnancy was 3.0% (410/13,457) (**Supplemental Table 1**) and increased from 0.5% in April 2020 to 10.5% in December 2021; October 2020 was the start of the positive slope (0.43 per month, p= <.01) (**Figure 1A)**. Trends among pregnant individuals living in urban areas were distinct from those living in rural areas (p=.02), with a steeper slope for urban residents during the first months of the pandemic, followed by a later increase among rural residents (**Figure 2A**). Trends in individuals with prenatal depression were flat followed by a steep increase after December 2020 (p<.01), in contrast to a more moderate steady increase among those without prenatal depression (**Figure 2B)**. Trends in individuals with GDM and HDP were similar to those in individuals without these conditions (**Supplemental Figure 1A-B)**.

**Figure 1:**
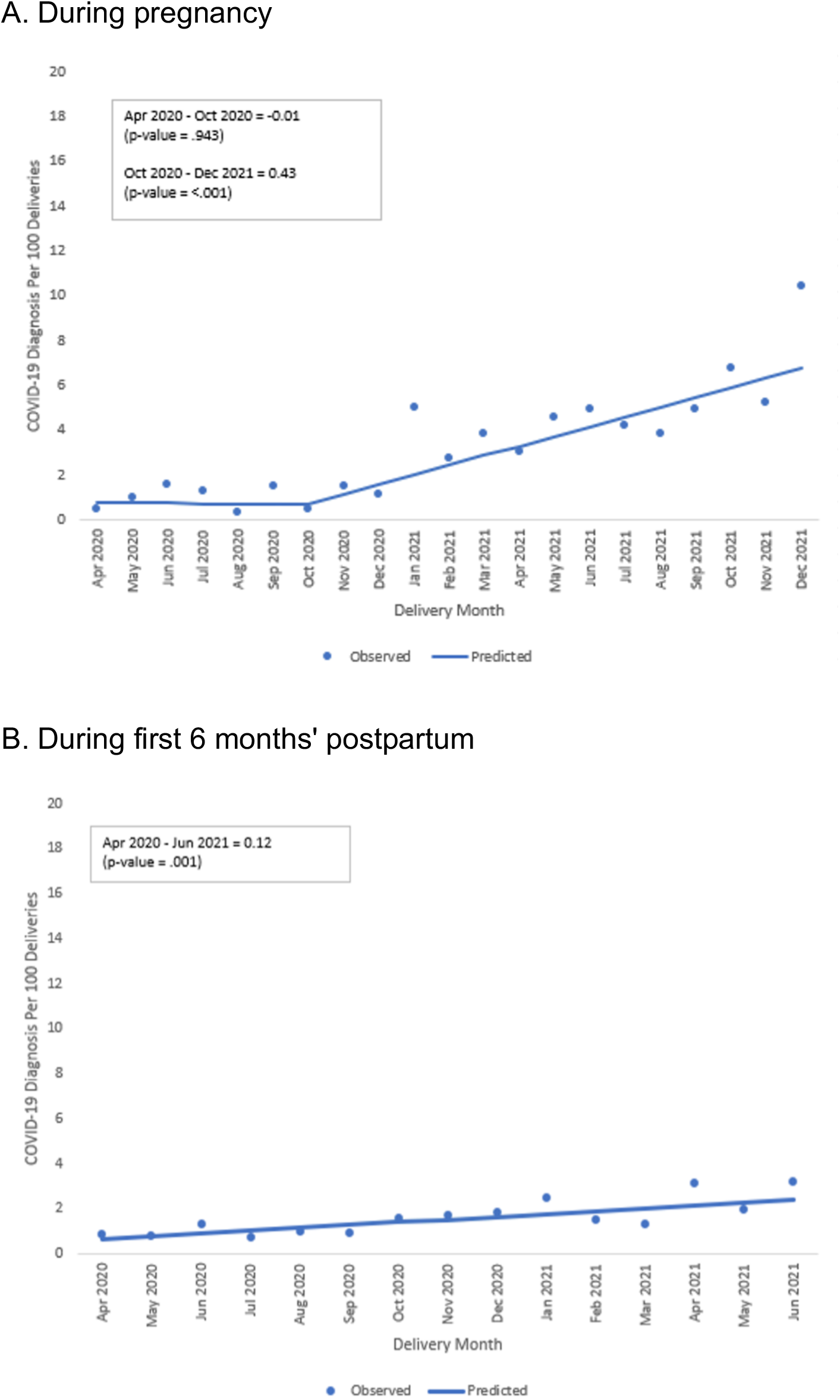
Prevalence of COVID-19 diagnosis by delivery month, Maine 2020-2021 Data source: Maine Health Data Organization’s All Payer Claims Data

**Figure 2:**
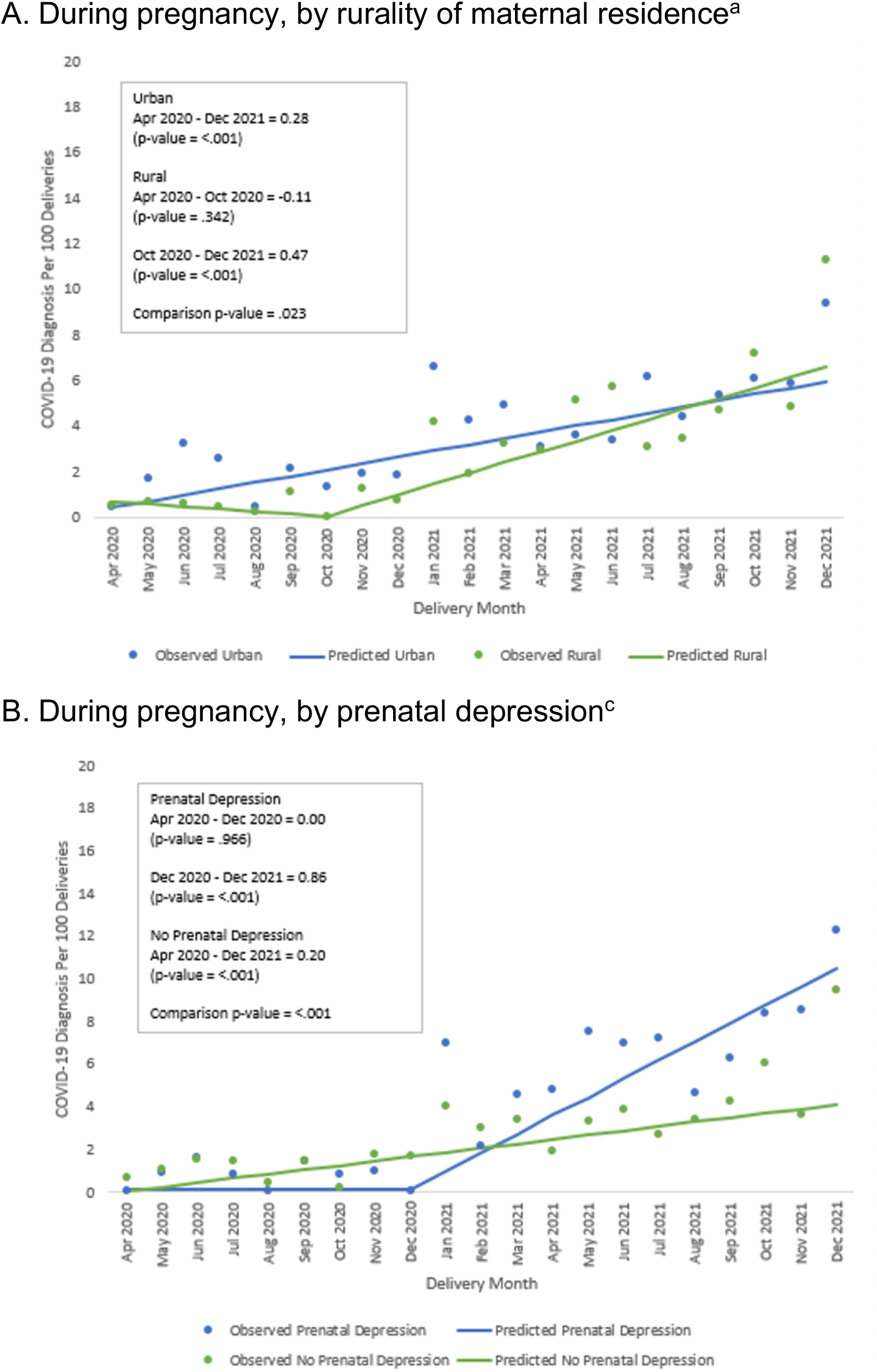
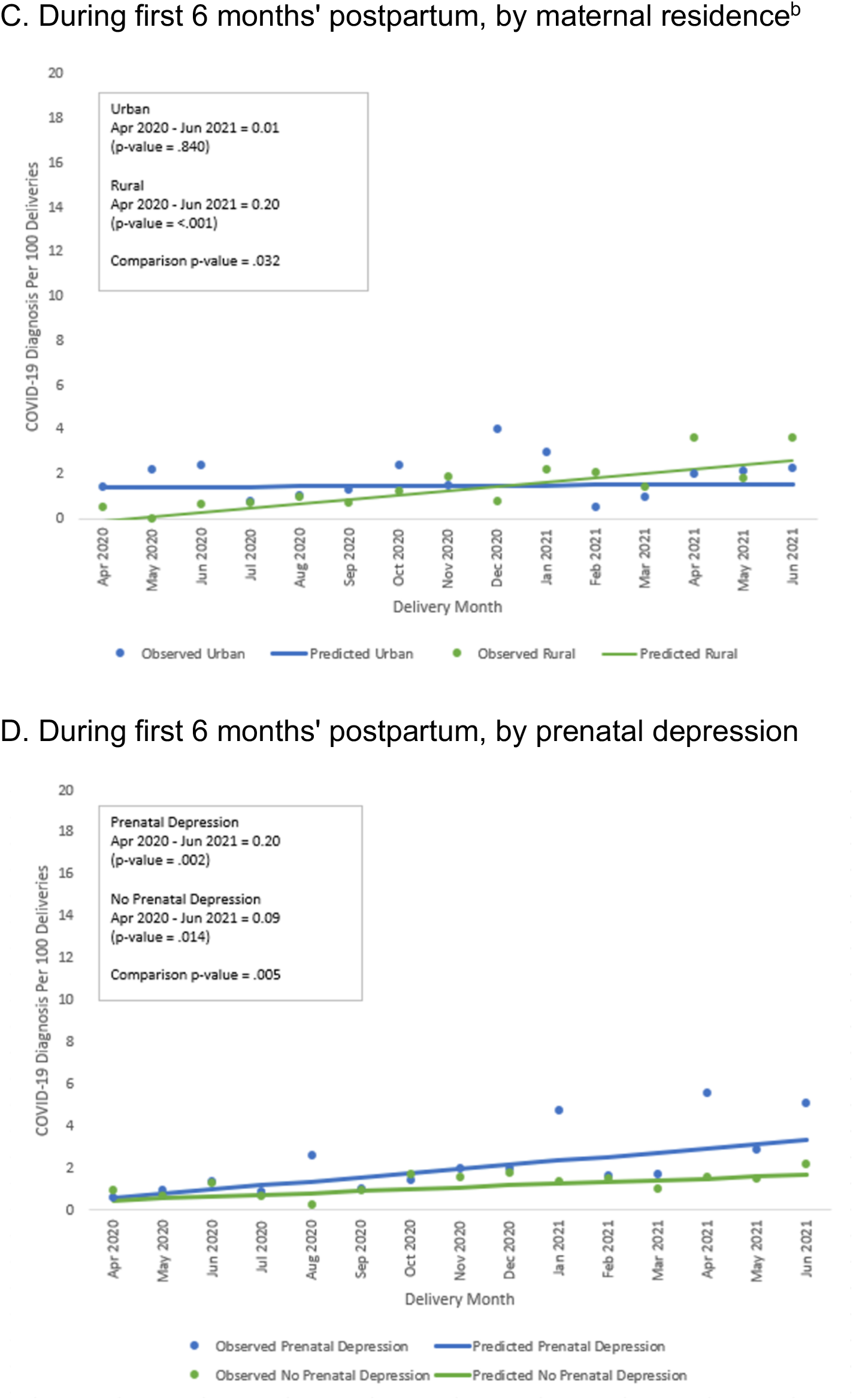
Prevalence of COVID-19 diagnosis by delivery month, rurality of maternal residence, and prenatal depression, Maine 2020-2021. Data source: Maine Health Data Organization’s All Payer Claims Data ^a^ Zero COVID-19 diagnoses for October 2020 replaced with 0.1 for rural residents. ^b^ Zero COVID-19 diagnoses for May 2020 replaced with 0.1 for rural residents. ^c^ Zero COVID-19 diagnoses for April 2020, August 2020, and December 2020 replaced with 0.1 for prenatal depression group.

### COVID-19 during the first 6 months’ postpartum

The prevalence of COVID-19 during the first 6 months’ postpartum was 1.6% (149/9,143) (**Supplemental Table 2**) and increased from 0.9% in April 2020 to 3.2% in June 2021 (slope = 0.12 per month, p<.01) **(Figure 1B)**. Trends among postpartum individuals living in urban areas were distinct from those living in rural areas (p=.03), with a steeper slope for rural residents over the course of the pandemic (**Figure 2C**). Trends in individuals with prenatal depression were steeper compared to those without prenatal depression (p<.01) (**Figure 2D**). Individuals without GDM and individuals without HDP had steeper increases in COVID-19 diagnoses postpartum compared to those with these conditions (p=.03 and p<.01, respectively) **(Supplemental Figure 1C-D)**.

## DISCUSSION

The prevalence of COVID-19 diagnosis among pregnant and postpartum individuals in Maine increased over the course of the pandemic. Our findings of COVID-19 diagnosis prevalence among pregnant individuals (3.0%) are similar to those reported from other studies with overlapping time periods (range 3.2% to 6.1%).^8,9^ In addition, we found trends in COVID-19 diagnosis among pregnant and postpartum individuals in Maine showed distinct patterns by rurality of residence and select pregnancy conditions.

Our study is the first to estimate prevalence of COVID-19 among pregnant and postpartum individuals for Maine, using data that captures both commercial and public insurance and examines differences by rurality, a known modifier of COVID-19 infections in the U.S.^3^ In addition, our study examined trends in COVID-19 diagnosis among individuals through the first 6 months’ postpartum, which has so far not been studied.

In terms of limitations, the introduction of universal COVID-19 testing upon admission for delivery hospitalizations in Maine may be responsible for some of the increases we found among pregnant individuals, but this date was not documented and likely varied by hospital. However, testing was presumably well underway by mid-2020,^10^ before we saw the start of the increase in October 2020. Additionally, our data did not capture any COVID-19 tests outside of the claims system (e.g. at-home tests); although, at-home tests weren’t widely used until after our study period ended.^11^ Lastly, claims data may provide an undercount for COVID-19 infections as compared with laboratory data.^12^

In conclusion, our results demonstrate the use of All Payer Claims Data for assessing perinatal trends in COVID-19 diagnosis. Findings can be used to inform interventions aimed at encouraging vaccination and preventing the spread of COVID-19 infections among pregnant and postpartum individuals.

## Data Availability

We used the Maine Health Data Organization's All Payer Claims Data as authorized under Data Request Number 2021040501.

https://mhdo.maine.gov/index.aspx

## ACKNOWLEDGMENTS

We thank the Maine Health Data Organization, which is responsible for the State of Maine’s All Payer Claims Data. We used the Maine Health Data Organization’s All Payer Claims Data as authorized under Data Request Number 2021040501.

## REFERENCES

1. Lassi ZS, Ana A, Das JK, et al. A systematic review and meta-analysis of data on pregnant women with confirmed COVID-19: Clinical presentation, and pregnancy and perinatal outcomes based on COVID-19 severity. J Glob Health. Jun 30 2021;11:05018. doi:10.7189/jogh.11.05018

2. Declercq E, Zephyrin L. Maternal Mortality in the United States: A Primer. Data Brief The Common Wealth Fund December 2020 2020;

3. Cuadros DF, Branscum AJ, Mukandavire Z, Miller FD, MacKinnon N. Dynamics of the COVID-19 epidemic in urban and rural areas in the United States. Ann Epidemiol. Jul 2021;59:16–20. doi:10.1016/j.annepidem.2021.04.007

4. Nation’s Urban and Rural Populations Shift Following 2020 Census. United States Census Bureau. Accessed April 11, 2023. https://www.census.gov/newsroom/press-releases/2022/urban-rural-populations.html

5. Governor Mills Welcomes End of State of Civil Emergency. State of Maine Office of Governor Janet T. Mills 2021. https://www.maine.gov/governor/mills/news/governor-mills-welcomes-end-state-civil-emergency-2021-06-30

6. Pfeiffer M, Gelsinger C, Palmsten K, Lipkind HS, Ackerman-Banks C, Ahrens KA. Rural-urban residence and sequelae of hypertensive disorders of pregnancy in the first year postpartum, 2007-2019. Pregnancy Hypertens. Feb 14 2023;32:10–17. doi:10.1016/j.preghy.2023.02.002

7. Data Warnings Displayed by Joinpoint. Accessed March 1, 2023. https://surveillance.cancer.gov/help/joinpoint/executing-joinpoint/input-data-2013-errors-and-warnings/data-warnings-displayed-by-joinpoint

8. Darling AM, Shephard H, Nestoridi E, Manning SE, Yazdy MM. SARS-CoV-2 infection during pregnancy and preterm birth in Massachusetts from March 2020 through March 2021. Paediatr Perinat Epidemiol. Feb 2023;37(2):93–103. doi:10.1111/ppe.12944

9. Steffen HA, Swartz SR, Jackson JB, et al. SARS-CoV-2 Infection during Pregnancy in a Rural Midwest All-delivery Cohort and Associated Maternal and Neonatal Outcomes. Am J Perinatol. May 2021;38(6):614–621. doi:10.1055/s-0041-1723938

10. Sutton D, Fuchs K, D’Alton M, Goffman D. Universal Screening for SARS-CoV-2 in Women Admitted for Delivery. N Engl J Med. May 28 2020;382(22):2163–2164. doi:10.1056/NEJMc2009316

11. Rader B, Gertz A AD. I, et al. Use of At-Home COVID-19 Tests — United States, August 23, 2021–March 12, 2022. MMWR Morb Mortal Wkly Rep. 2022;71:489–494. doi:http://dx.doi.org/10.15585/mmwr.mm7113e1

12. Regan AK, Arah OA, Sullivan SG. Performance of diagnostic coding and laboratory testing results to measure COVID-19 during pregnancy and associations with pregnancy outcomes. Paediatr Perinat Epidemiol. Jul 2022;36(4):508–517. doi:10.1111/ppe.12863

