## Supplemental Materials for "COVID-19 Prevalence and Trends Among Pregnant and Postpartum Individuals in Maine by Rurality and Pregnancy Conditions"

**Supplemental Table 1.** Maternal and pregnancy characteristics, by COVID-19 diagnosis during pregnancy in Maine, 2020-2021

| | TOTAL | | No Covid-19 diagnosis n=13047 | Covid-19 diagnosis n=410 |
|---|---|---|---|---|
| | N | % | % | % |
| Total | 13457 | 100.0 | 100.0 | 100.0 |
| %, row | | | 97.0 | 3.0 |
| Maternal age at delivery, years | | | | |
| Missing | 2 | 0.1 | 0.0 | 0.0 |
| 15 to 19 | 365 | 2.7 | 2.7 | [a] |
| 20 to 24 | 2379 | 17.7 | 17.6 | 21.0 |
| 25 to 29 | 3701 | 27.5 | 27.4 | 31.2 |
| 30 to 34 | 4237 | 31.5 | 31.6 | 29.0 |
| 35+ | 2773 | 20.6 | 20.7 | 16.3 |
| Cesarean section | | | | |
| Yes | 4033 | 30.0 | 29.9 | 31.2 |
| Gestational age | | | | |
| At least 37 weeks | 12225 | 90.8 | 90.9 | 89.5 |
| 20 to <37 weeks | 1232 | 9.2 | 9.1 | 10.5 |
| Insurance coverage | | | | |
| Medicaid | 7669 | 57.0 | 56.8 | 62.4 |
| Private | 5772 | 42.9 | 43.1 | 37.6 |
| Medicare | 16 | 0.1 | 0.1 | [a] |
| Rurality | | | | |
| Metro | 4770 | 35.5 | 35.2 | 42.2 |
| Large Rural | 4951 | 36.8 | 36.9 | 33.2 |
| Small Rural | 1829 | 13.6 | 13.6 | 13.7 |
| Isolated Rural | 1907 | 14.2 | 14.3 | 11.0 |
| Loss of health insurance postpartum | | | | |
| 0 to 5 months | 4314 | 32.1 | 31.5 | 50.5 |
| 6 to 11 months | 3859 | 28.7 | 28.4 | 36.6 |
| 12 to 23 months | 5284 | 39.3 | 40.1 | 12.9 |
| Prenatal depression | | | | |
| Yes | 4459 | 32.9 | 32.6 | 42.7 |
| Hypertensive disorders of pregnancy | | | | |
| Yes | 2537 | 18.9 | 18.7 | 23.4 |
| Gestational diabetes | | | | |
| Yes | 1531 | 11.4 | 11.3 | 12.4 |

Data source: Maine Health Data Organization's All Payer Claims Data
[a] Suppressed cell count per data use agreement with Maine Health Data Organization

**Supplemental Table 2.** Maternal and pregnancy characteristics, by Covid-19 diagnosis during the first 6 months' postpartum in Maine, 2020-2021

| | TOTAL | | No Covid-19 diagnosis n=8994 | Covid-19 diagnosis n=149 |
|---|---|---|---|---|
| | N | % | % | % |
| Total | 9143 | 100.0 | 100.0 | 100.0 |
| %, row | | | 98.4 | 1.6 |
| Maternal age at delivery, years | | | | |
| Missing | 2 | 0.0 | 0.0 | 0.0 |
| 15 to 19 | 260 | 2.8 | 2.9 | [a] |
| 20 to 24 | 1619 | 17.7 | 17.6 | 23.5 |
| 25 to 29 | 2543 | 27.8 | 27.8 | 26.2 |
| 30 to 34 | 2903 | 31.8 | 31.8 | 28.9 |
| 35+ | 1816 | 19.9 | 19.9 | 18.8 |
| Cesarean section | | | | |
| Yes | 2794 | 30.6 | 30.5 | 32.9 |
| Gestational age | | | | |
| At least 37 weeks | 8295 | 90.7 | 90.8 | 87.9 |
| 20 to <37 weeks | 848 | 9.3 | 9.2 | 12.1 |
| Insurance coverage | | | | |
| Medicaid | 5415 | 59.2 | 59.0 | 72.5 |
| Private | 3715 | 40.6 | 40.9 | 26.9 |
| Medicare | 13 | 0.1 | 0.1 | [a] |
| Rurality | | | | |
| Metro | 3177 | 34.8 | 34.7 | 40.3 |
| Large Rural | 3340 | 36.5 | 36.6 | 32.9 |
| Small Rural | 1278 | 14.0 | 14.0 | 11.4 |
| Isolated Rural | 1348 | 14.7 | 14.7 | 15.4 |
| Loss of health insurance postpartum | | | | |
| 6 to 11 months | 3859 | 42.2 | 42.0 | 57.1 |
| 12 to 23 months | 5284 | 57.8 | 58.0 | 43.0 |
| Prenatal depression | | | | |
| Yes | 3068 | 33.6 | 33.3 | 48.3 |
| Hypertensive disorders of pregnancy | | | | |
| Yes | 1738 | 19.0 | 18.9 | 24.2 |
| Gestational diabetes | | | | |
| Yes | 1072 | 11.7 | 11.7 | 11.4 |

Data source: Maine Health Data Organization's All Payer Claims Data
[a] Suppressed cell count per data use agreement with Maine Health Data Organization

**Supplemental Figure 1**: Prevalence of COVID-19 diagnosis by delivery month, hypertensive disorders of pregnancy, and gestational diabetes, Maine 2020-2021

A. During pregnancy, by hypertensive disorder of pregnancy[a]

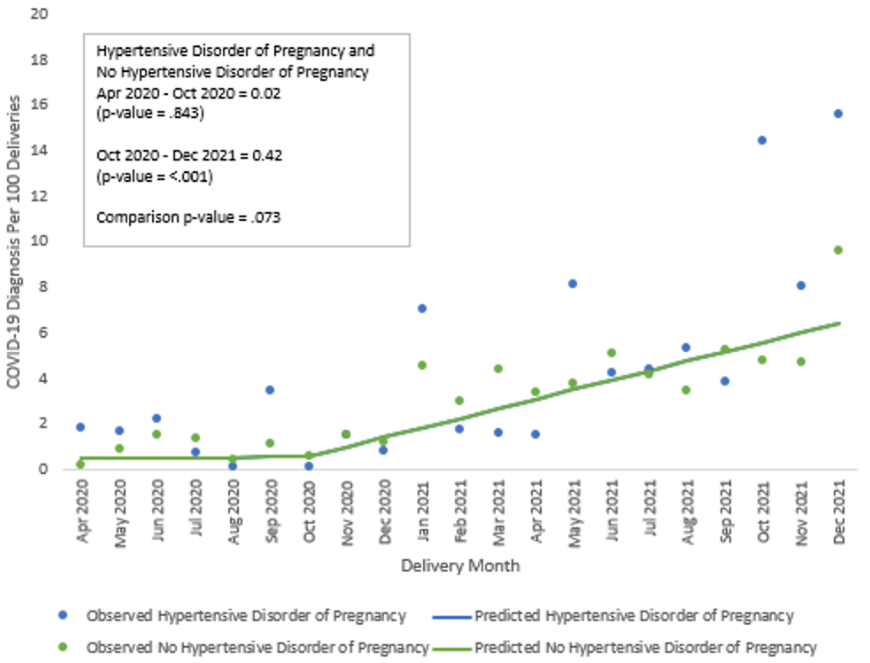


B. During pregnancy, by gestational diabetes[b]

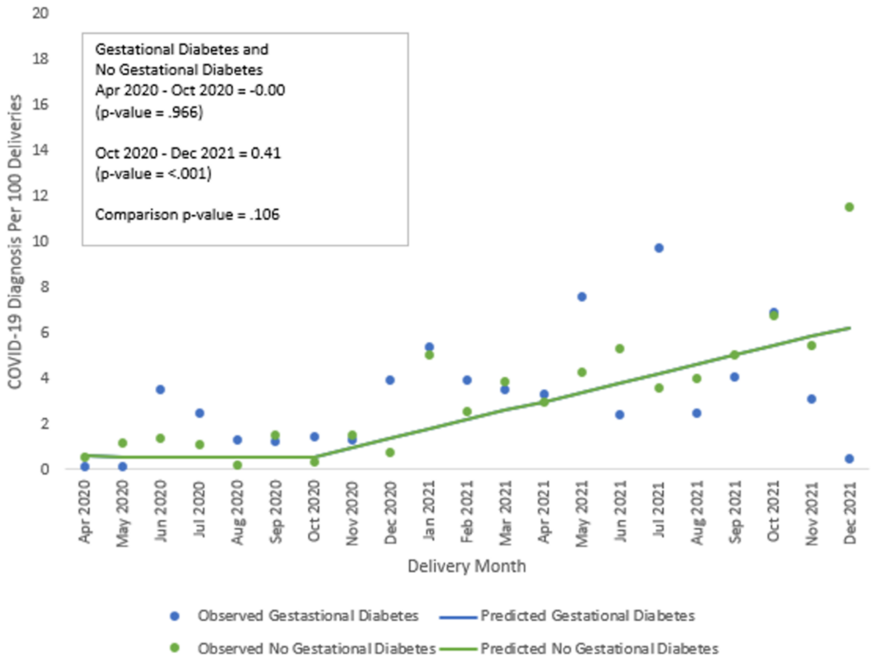

C. During first 6 months' postpartum, by hypertensive disorder of pregnancy[c]

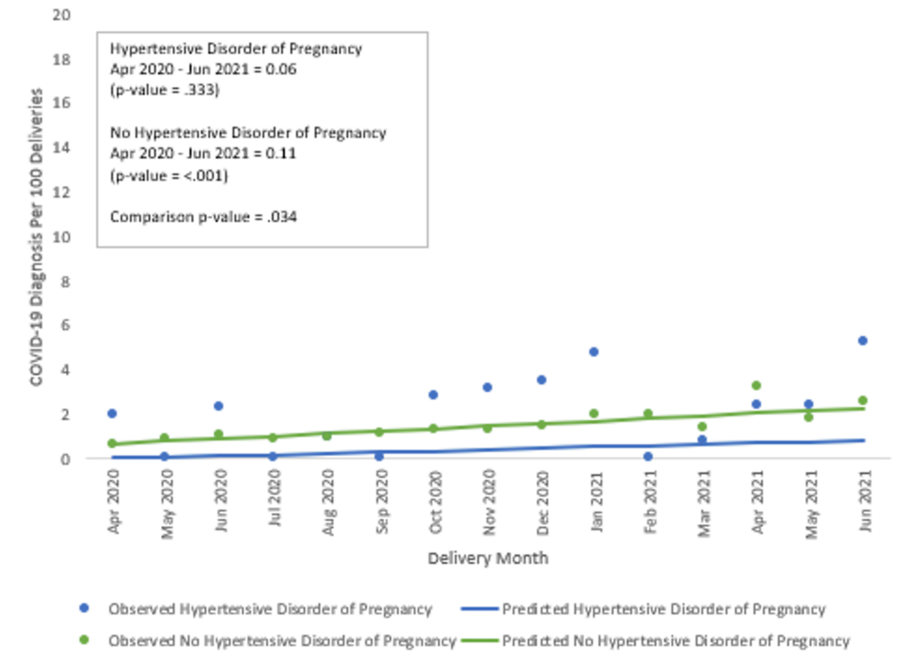


D. During first 6 months' postpartum, by gestational diabetes[d]

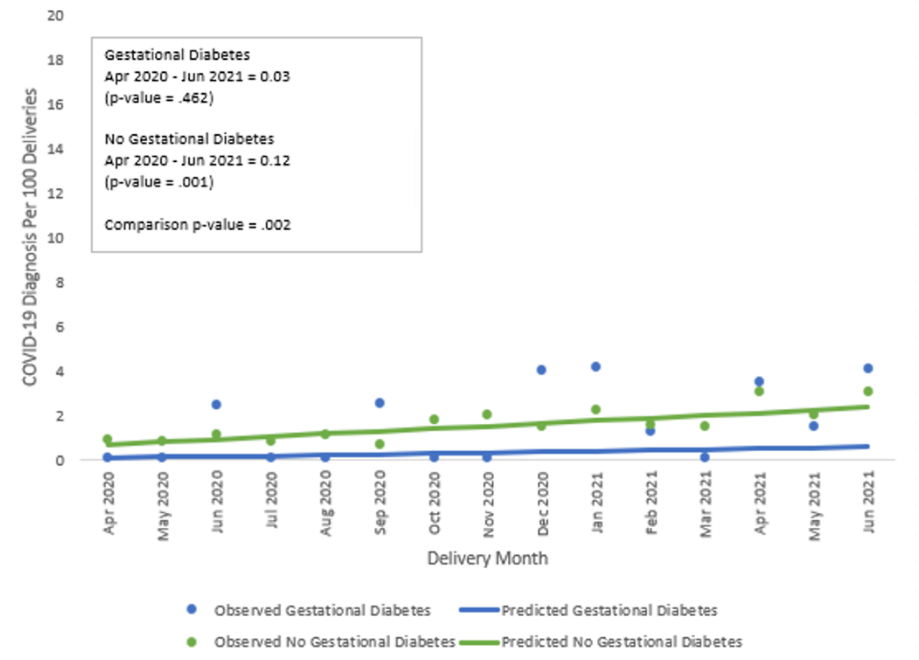


Data source: Maine Health Data Organization's All Payer Claims Data
[a] Zero COVID-19 diagnoses for August 2020 and October 2020 replaced with 0.1 for HDP. Trend line for individuals with hypertensive disorders of pregnancy (HDP) completely overlaps and obscures trend line for individuals without HDP.

[b] Zero COVID-19 diagnoses for April 2020, May 2020 and December 2021 replaced with 0.1 for GDM. Trend line for individuals with gestational diabetes (GDM) completely overlaps and obscures trend line for individuals without GDM.

[c] Zero COVID-19 diagnoses for May 2020, July 2020, September 2020, and February 2021 replaced with 0.1 for HDP.

[d] Zero COVID-19 diagnoses for April 2020, May 2020, July 2020, August 2020, October 2020, November 2020, and March 2021 replaced with 0.1 for GDM.
